# Regional performance variation in external validation of four prediction models for severity of COVID-19 at hospital admission: An observational multi-centre cohort study

**DOI:** 10.1101/2021.03.26.21254390

**Authors:** Kristin E. Wickstrøm, Valeria Vitelli, Ewan Carr, Aleksander R. Holten, Rebecca Bendayan, Andrew H. Reiner, Daniel Bean, Tom Searle, Anthony Shek, Zeljko Kraljevic, James Teo, Richard Dobson, Kristian Tonby, Alvaro Köhn- Luque, Erik K. Amundsen

## Abstract

**Background:** Several prediction models for coronavirus disease-19 (COVID-19) have been published. Prediction models should be externally validated to assess their performance before implementation. This observational cohort study aimed to validate published models of severity for hospitalized patients with COVID-19 using clinical and laboratory predictors.

**Methods:** Prediction models fitting relevant inclusion criteria were chosen for validation. The outcome was either mortality or a composite outcome of mortality and ICU admission (severe disease). 1295 patients admitted with symptoms of COVID-19 at Kings Cross Hospital (KCH) in London, United Kingdom, and 307 patients at Oslo University Hospital (OUH) in Oslo, Norway were included. The performance of the models was assessed in terms of discrimination and calibration.

**Results:** We identified two models for prediction of mortality (referred to as Xie and Zhang1) and two models for prediction of severe disease (Allenbach and Zhang2).

The performance of the models was variable. For prediction of mortality Xie had good discrimination at OUH with an area under the receiver-operating characteristic (AUROC) 0.87 [95 % confidence interval (CI) 0.79-0.95] and acceptable discrimination at KCH, AUROC 0.79 [0.76-0.82]. In prediction of severe disease, Allenbach had acceptable discrimination (OUH AUROC 0.81 [0.74-0.88] and KCH AUROC 0.72 [0.68-0.75]). The Zhang models had moderate to poor discrimination. Initial calibration was poor for all models but improved with recalibration.

**Conclusions:** The performance of the four prediction models was variable. The Xie model had the best discrimination for mortality, while the Allenbach model had acceptable results for prediction of severe disease.

## Introduction

Severe acute respiratory syndrome coronavirus 2 (SARS-CoV-2) was discovered in Wuhan, China in December 2019. The virus was shown to cause viral pneumonia, later designated as coronavirus disease 2019 (COVID-19) [1]. The disease has evolved as a pandemic with an extensive amount of severe cases with high mortality [2]. Several biomarkers, clinical and epidemiological parameters have been associated with disease severity [3, 4]. Practical tools for prediction of prognosis in COVID-19 patients are still lacking in clinical practice [5, 6]. Prediction models can be crucial to prioritize patients needing hospitalization, intensive care treatment, or future individualized therapy.

Since the onset of the pandemic, the number of prediction models for COVID-19 patients has been continuously growing [7]. Prediction models should be validated in different populations with a sufficient number of patients reaching the outcome before implementation [8-10]. A validation study of 22 prediction models at one site was recently published [6]. Interestingly, this study found that none of the models performed better than oxygen saturation alone, even though the performance at the original study sites in most cases was much better.

This study aimed to validate published prediction models of severity and mortality for hospitalized patients based on laboratory and clinical values in COVID-19 cohorts from London (United Kingdom) and Oslo (Norway).

The study is reported according to the guidelines in “Transparent reporting of a multivariable prediction model for individual prognosis or diagnosis” (TRIPOD) [11] and has also followed recommendations from “Prediction Model Risk of Bias Assessment Tool” (PROBAST) [12].

## Methods

### Selection of prediction models

A literature search was performed to select prediction models for validation. Published articles or preprint manuscripts were included until 29.05.2020. A structured search was performed in PubMed with the words “COVID-19” and “prediction model” or “machine learning” or “prognosis model”. Prediction models included in the review by Wynants et. al. [7] in May 2020 were also investigated, as well as search for articles/preprints citing Wynants et. al. using Google Scholar 18.05.2020.

The inclusion criteria for selection of multivariable prediction models were: (1) Symptomatic hospitalized patients over 18 years with PCR confirmed COVID-19; (2) outcomes including respiratory failure or intensive care unit (ICU) admission or death or composite outcomes of these. (3) The predictive models had to include at least one laboratory test as we wanted to explore models that combined clinical and laboratory variables (4). All variables had to be available in the OUH dataset and the model had to be described in adequate detail.

### Study design and participants

The study was performed as a retrospective validation study with adult patients hospitalized with COVID-19. Two cohorts were included: (1) Oslo University Hospital (OUH) in Norway, (2) Kings Cross Hospital (KCH) in London, United Kingdom. The patients included were all adult inpatients testing positive for SARS-CoV-2 by real-time polymerase chain reaction (RT-PCR) with symptoms consistent with COVID-19 at admission. SARS-CoV-2 -positive patients admitted for conditions not related to COVID-19 were excluded, e.g. pregnancy-related conditions or trauma. Patients referred from other hospitals were also excluded, as we did not have access to measurements from the first hospital admission.

### OUH cohort

OUH is a large urban university hospital. Patients admitted between 6^th^ March and 31^th^ December 2020 were included. The OUH project protocol was approved by the Regional Ethical Committee of South East Norway (Reference 137045). All patients with confirmed COVID-19 were included in the quality registry “COVID19 OUS”, approved by the data protection officer (Reference 20/08822). Informed consent was waived because of the strictly observational nature of the project. Demographics, clinical variables and hospital stay information were manually recorded in the registry and merged with laboratory results exported from the laboratory information system in Microsoft Excel.

### KCH cohort

In the KCH cohort patients were admitted between 23^rd^ February to 1^st^ May 2020 at two hospitals (King’s College Hospital and Princess Royal University Hospital) in South East London (UK) of Kings College Hospital NHS Foundation Trust.

Data (demographics, emergency department letters, discharge summaries, lab results, vital signs) were retrieved from components of the electronic health record (EHR) using a variety of natural language processing (NLP) informatics tools belonging to the CogStack ecosystem [13]. The project operated under London South East Research Ethics Committee (reference 18/LO/2048) approval granted to the King’s Electronic Records Research Interface (KERRI); specific work on COVID-19 research was reviewed with expert patient input on a virtual committee with Caldicott Guardian oversight. Data from this cohort has been published in prior studies [14, 15].

### Missing values

Predictive variables were collected from the admission to the emergency department (ED). If not available in the ED, the first available values within 24 hours from hospital admission were used. Missing values (i.e. no recorded values within 24 hours) were generally imputed using k-nearest neighbors (KNN) although we tested more advanced techniques based on Python’s scikit-learn IterativeImputer, including random forest-based imputation, and multiple imputation using Bayesian ridge and Gaussian process methods [16, 17].

### Statistical analyses and performance measurements for the prediction models

Univariate comparisons between patients with ‘mild’ versus ‘severe’ disease were carried out for continuous (Wilcoxon rank-sum test) and binary (*X*^*2*^ test) measures. Severe disease was defined as transfer to ICU or in-hospital mortality.

Validation of the selected prediction models was assessed with discrimination and calibration as recommended in TRIPOD [11]. Discrimination is the ability of the model to differentiate between those who do or do not experience the outcome. It is commonly estimated by concordance index (c-index) which is identical to the area under the receiver-operating characteristic curve (AUROC) for models with binary endpoints. The discrimination for the models at OUH and KCH was also compared to the discrimination in the original development cohort and to the external validation by Gupta et. al [6]. Calibration is the agreement between the observed outcomes and the outcome predictions from the model. It is preferably reported by a calibration plot, intercept and slope.

Models were recalibrated by adjusting the intercept of the logistic regression models according to the frequency of outcomes at each study site [18]. All statistical analyses were conducted in Python 3.7 and R 3.4 [19].

## Results

### Selection of prediction models

Four publications comprising five prediction models fit our inclusion criteria [14, 20-22]. The inclusion process is illustrated in Figure 1. However, since one of the models was developed at KCH and validated at OUH in a previous publication [14], only four models are presented here. The four models are referred to as ‘Xie’[20], ‘Zhang1’, ‘Zhang2’[21] and ‘Allenbach’[22].

**Figure 1:**
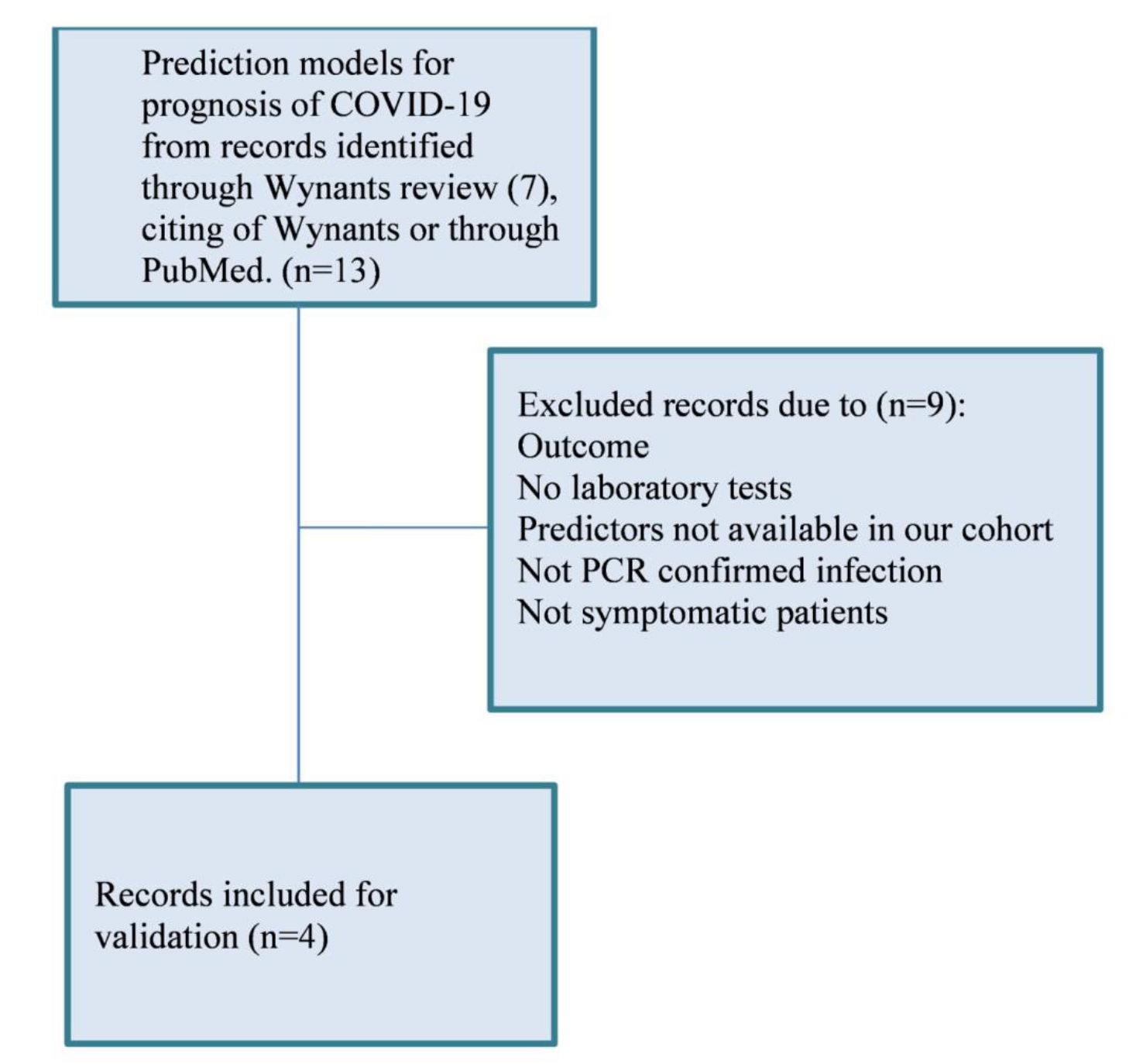
Selection of prediction models for validation.

Information on the predictor variables and outcomes of the four models are summarized in Table 1.

**Table 1:** Predictors and outcomes in the three prediction models.

|  | **Zhang models** | **Xie model** | **Allenbach model** |
| --- | --- | --- | --- |
| Country of development cohort | China | China | France |
| Predictors | Age, sex, neutrophil count, lymphocyte count, platelets count, CRP and creatinine at admission. | Age, LDH, SpO2, lymphocyte count (log, due to extreme value). | CRP (per 100mg/L), age, lymphocyte count, WHO scale (22) by admission. |
| Outcome | 1. Mortality<br>2. Poor outcome, defined as developing ARDS, receiving intubation or ECMO treatment, ICU admission and death. | 1. Hospital mortality | 1. ICU transfer or death by 14 days after admission. |

CRP; C-reactive protein, LDH; lactate dehydrogenase, SpO2; Peripheral oxygen saturation, WHO; World Health Organization, ICU; Intensive care unit, NEWS2; National Early Warning score 2, eGFR; estimated glomerular filtration rate, ECMO; extracorporeal membrane oxygenation, ARDS; acute respiratory distress syndrome.

All predictors were measured at hospital admission. Missing values and imputation methods used in the development cohorts were not well described. The Xie model had hospital mortality as the only outcome. Zhang presented two models with different outcomes: (1) Mortality and (2) Composite outcome of mortality or ‘poor outcome’. Poor outcome was defined as acute respiratory distress syndrome (ARDS), intubation or extracorporeal membrane oxygenation (ECMO) treatment, ICU admission or death. The Allenbach model used a composite outcome of transfer to ICU or mortality within 14 days of hospital admission. There were no details of the censoring date in the original studies. Mortality during the hospital stay was used for the OUH cohort and for the KCH cohort hospital mortality at data collection time.

All prediction models were based on multiple logistic regression and presented coefficients and intercepts for the different variables that enabled the calculation of risk prediction for our cohorts. Allenbach additionally provided an 8-point scoring system derived from the logistic regression model. However, we chose to use the regression model for calculation as this retains as much information as possible.

### Description of the cohorts

Patient characteristics for the three development cohorts and the KCH and OUH cohorts are shown in Table S1 (supplementary material).

Since the three models use different outcomes and timeframes, the number of patients included in each validation is not the same. An overview of missing values is presented in Table 2. Missing values were imputed via simple imputation and multiple imputations [17]. Preliminary analyses showed no differences between AUROCs calculated with different imputation methods (see Table S2). Thus, the simple imputation method k-nearest neighbor was used for the rest of this paper. At KCH the number of missing values was very high for LDH (87.8 %) and relatively high for SpO2 (33.3 %) and WHO scale (33.8 %).

**Table 2:**
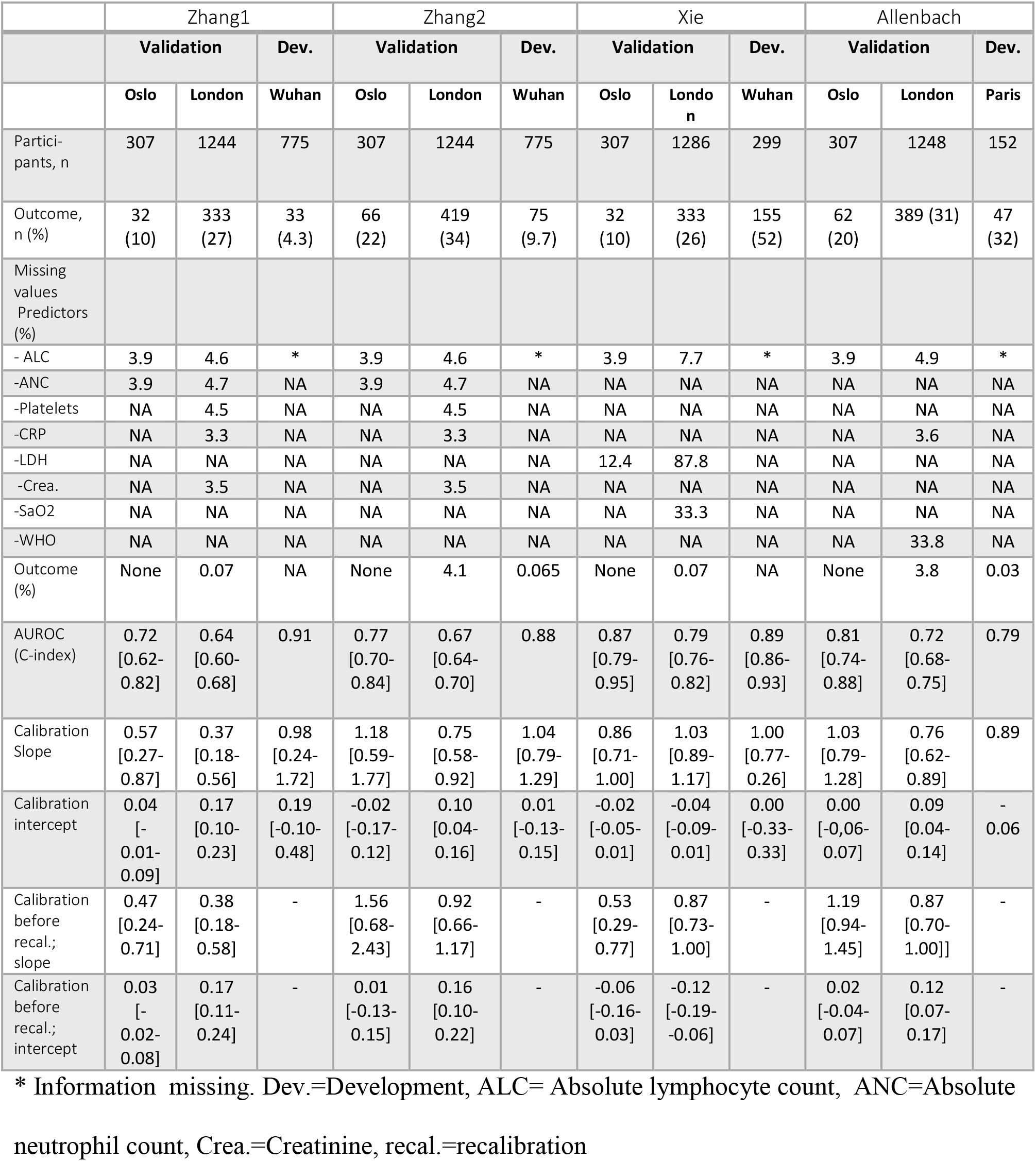
Missing values, and results for discrimination and calibration.

The OUH cohort consisted of 307 patients while the KCH cohort consisted of 1295 patients (Figure S1). For the OUH cohort median age was 60 years with 57 % males, while in the KCH cohort the median age was 69 with 59 % males. In the OUH cohort, 32 patients died in the hospital (10.4 %), while 333 (26.8 %) had died at the hospital by data collection time in the KCH cohort. For the composite outcome death or ICU transfer, the number of patients with the outcome was 66 (21.5 %) at OUH and 419 (33.7 %) at KCH.

The percentage of patients with hypertension and diabetes was higher in the KCH cohort (54 % and 35 %, respectively) than in the OUH cohort (34 % and 21 %, respectively). The patients at KCH also had higher levels of CRP, creatinine, LDH, and possibly a lower number of lymphocytes than the OUH patients; all of which are known predictors for severe COVID-19.

In Table 3, univariate associations are presented for mild/moderate and severe groups for the KCH and OUH cohorts. In general, the same variables were predictive for severe disease at KCH and OUH; except for ischemic heart disease, temperature and platelets which were associated with severe disease at OUH, but not KCH.

**Table 3:**
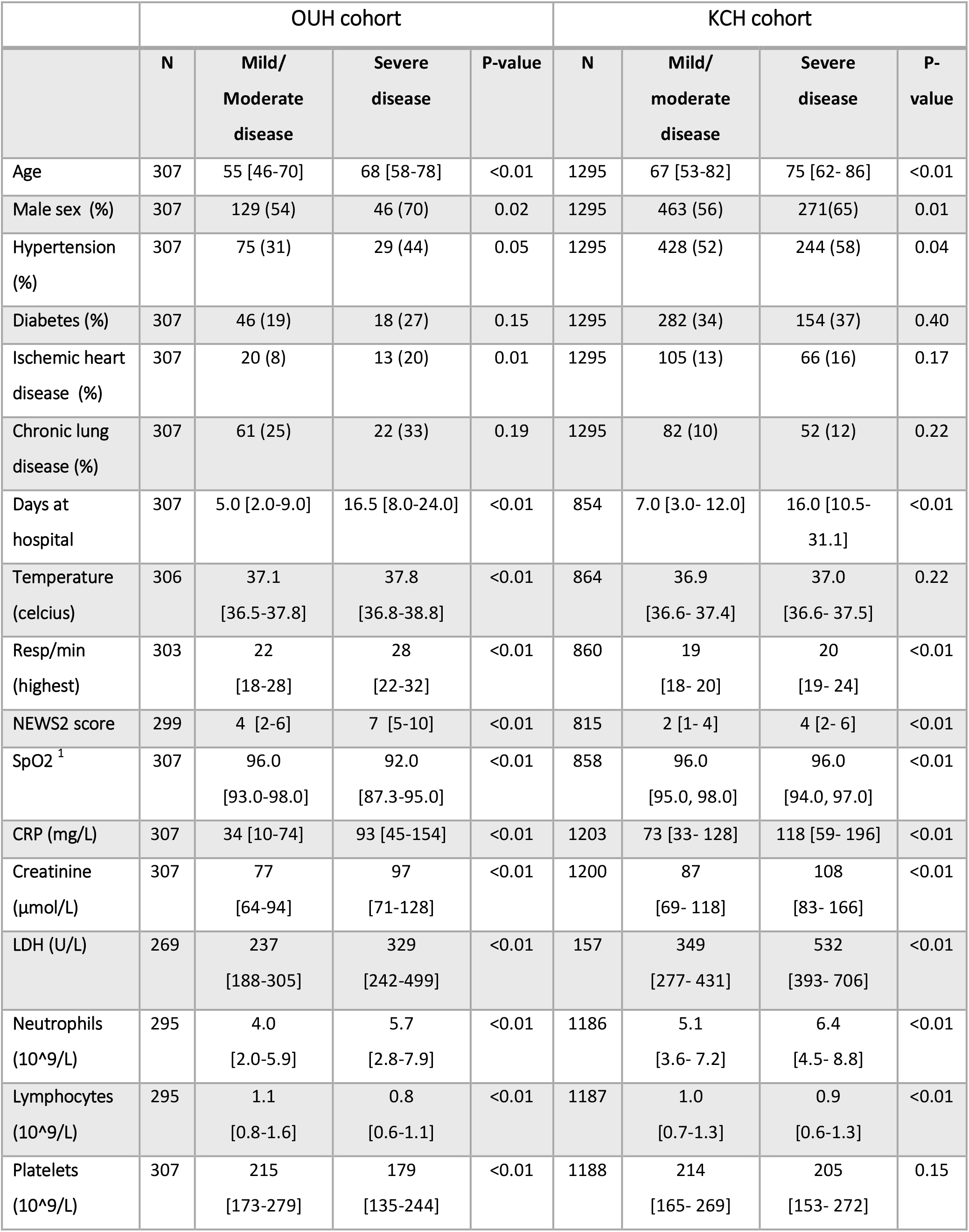
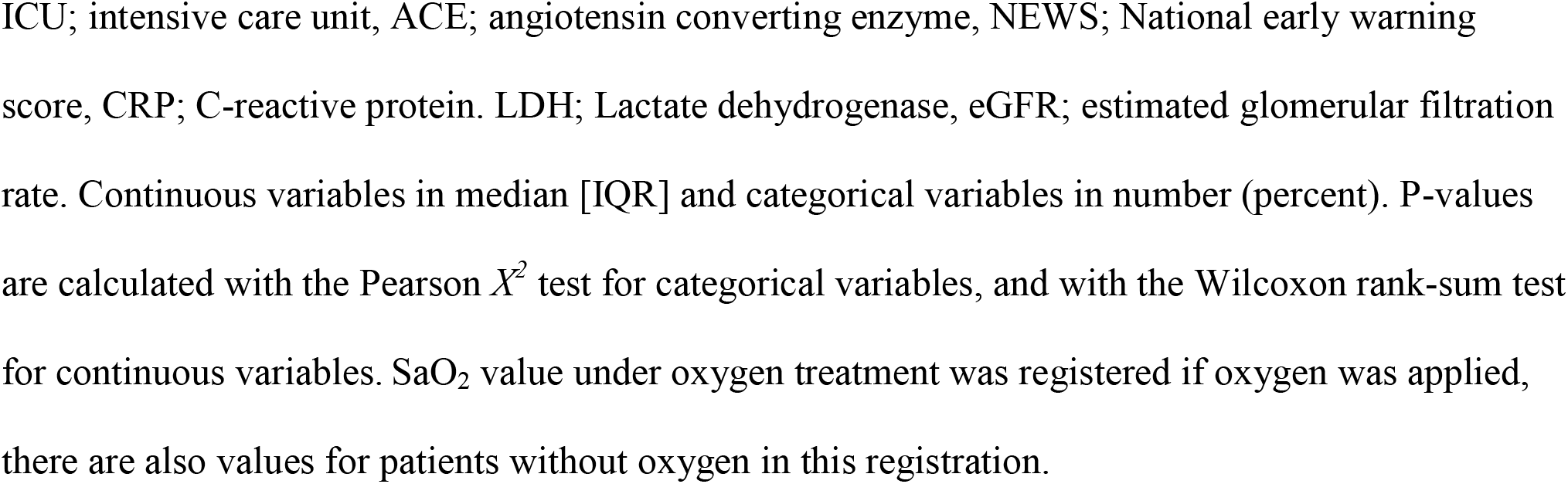
Univariate analysis of predictors at KCH and OUH.

|  | OUH cohort |  |  |  | KCH cohort |  |  |  |
| --- | --- | --- | --- | --- | --- | --- | --- | --- |
|  | N | Mild/<br>Moderate<br>disease | Severe<br>disease | P-value | N | Mild/<br>moderate<br>disease | Severe<br>disease | P-value |
| Age | 307 | 55 [46-70] | 68 [58-78] | <0.01 | 1295 | 67 [53-82] | 75 [62- 86] | <0.01 |
| Male sex (%) | 307 | 129 (54) | 46 (70) | 0.02 | 1295 | 463 (56) | 271(65) | 0.01 |
| Hypertension (%) | 307 | 75 (31) | 29 (44) | 0.05 | 1295 | 428 (52) | 244 (58) | 0.04 |
| Diabetes (%) | 307 | 46 (19) | 18 (27) | 0.15 | 1295 | 282 (34) | 154 (37) | 0.40 |
| Ischemic heart disease (%) | 307 | 20 (8) | 13 (20) | 0.01 | 1295 | 105 (13) | 66 (16) | 0.17 |
| Chronic lung disease (%) | 307 | 61 (25) | 22 (33) | 0.19 | 1295 | 82 (10) | 52 (12) | 0.22 |
| Days at hospital | 307 | 5.0 [2.0-9.0] | 16.5 [8.0-24.0] | <0.01 | 854 | 7.0 [3.0- 12.0] | 16.0 [10.5- 31.1] | <0.01 |
| Temperature (celcius) | 306 | 37.1<br>[36.5-37.8] | 37.8<br>[36.8-38.8] | <0.01 | 864 | 36.9<br>[36.6- 37.4] | 37.0<br>[36.6- 37.5] | 0.22 |
| Resp/min (highest) | 303 | 22<br>[18-28] | 28<br>[22-32] | <0.01 | 860 | 19<br>[18- 20] | 20<br>[19- 24] | <0.01 |
| NEWS2 score | 299 | 4 [2-6] | 7 [5-10] | <0.01 | 815 | 2 [1- 4] | 4 [2- 6] | <0.01 |
| SpO2 <sup>1</sup> | 307 | 96.0<br>[93.0-98.0] | 92.0<br>[87.3-95.0] | <0.01 | 858 | 96.0<br>[95.0, 98.0] | 96.0<br>[94.0, 97.0] | <0.01 |
| CRP (mg/L) | 307 | 34 [10-74] | 93 [45-154] | <0.01 | 1203 | 73 [33- 128] | 118 [59- 196] | <0.01 |
| Creatinine (μmol/L) | 307 | 77<br>[64-94] | 97<br>[71-128] | <0.01 | 1200 | 87<br>[69- 118] | 108<br>[83- 166] | <0.01 |
| LDH (U/L) | 269 | 237<br>[188-305] | 329<br>[242-499] | <0.01 | 157 | 349<br>[277- 431] | 532<br>[393- 706] | <0.01 |
| Neutrophils (10<sup>9</sup>/L) | 295 | 4.0<br>[2.0-5.9] | 5.7<br>[2.8-7.9] | <0.01 | 1186 | 5.1<br>[3.6- 7.2] | 6.4<br>[4.5- 8.8] | <0.01 |
| Lymphocytes (10<sup>9</sup>/L) | 295 | 1.1<br>[0.8-1.6] | 0.8<br>[0.6-1.1] | <0.01 | 1187 | 1.0<br>[0.7-1.3] | 0.9<br>[0.6-1.3] | <0.01 |
| Platelets (10<sup>9</sup>/L) | 307 | 215<br>[173-279] | 179<br>[135-244] | <0.01 | 1188 | 214<br>[165- 269] | 205<br>[153- 272] | 0.15 |

ICU; intensive care unit, ACE; angiotensin converting enzyme, NEWS; National early warning score, CRP; C-reactive protein. LDH; Lactate dehydrogenase, eGFR; estimated glomerular filtration rate. Continuous variables in median [IQR] and categorical variables in number (percent). P-values are calculated with the Pearson $\chi^2$ test for categorical variables, and with the Wilcoxon rank-sum test for continuous variables. SaO<sub>2</sub> value under oxygen treatment was registered if oxygen was applied, there are also values for patients without oxygen in this registration.

### Performance of the prediction models

The validation of the four prediction models with both the OUH and KCH cohorts is presented in terms of discrimination (AUROC) and calibration (slope and intercept) in Table 2 and Figures 2 and 3, respectively. For the models predicting mortality, the Xie model had the highest AUROC both in the KCH cohort (0.79; 95 % CI 0.76-0.82) and the OUH cohort (0.87; 95 % CI 0.79-0.95). The Zhang1 model had a lower AUROC at both KHC (0.64; 95 % CI 0.60-0.68) and OUH (0.72; 95 % CI 0.62-0.82).

**Figure 2:**
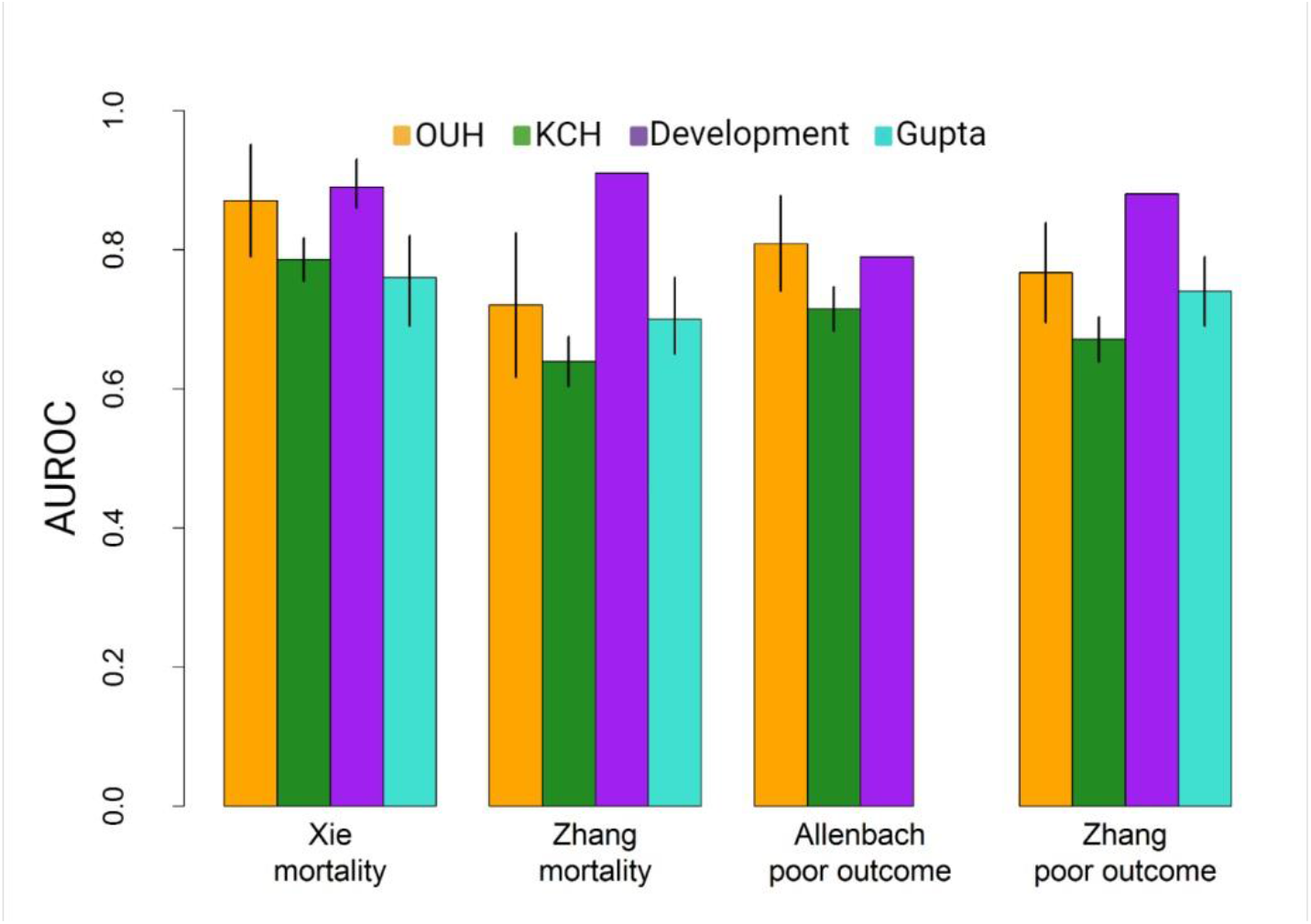
AUROCs from validation of the four models at the KCH and OUH cohorts, and the original AUROC from development cohorts [20-22]. Also shown are the results from the external validation of the Xie and Zhang models by Gupta et al [6]. Lines represent the 95% CIs of the AUROCs. For the development cohorts only Xie reported confidence intervals.

**Figure 3:**
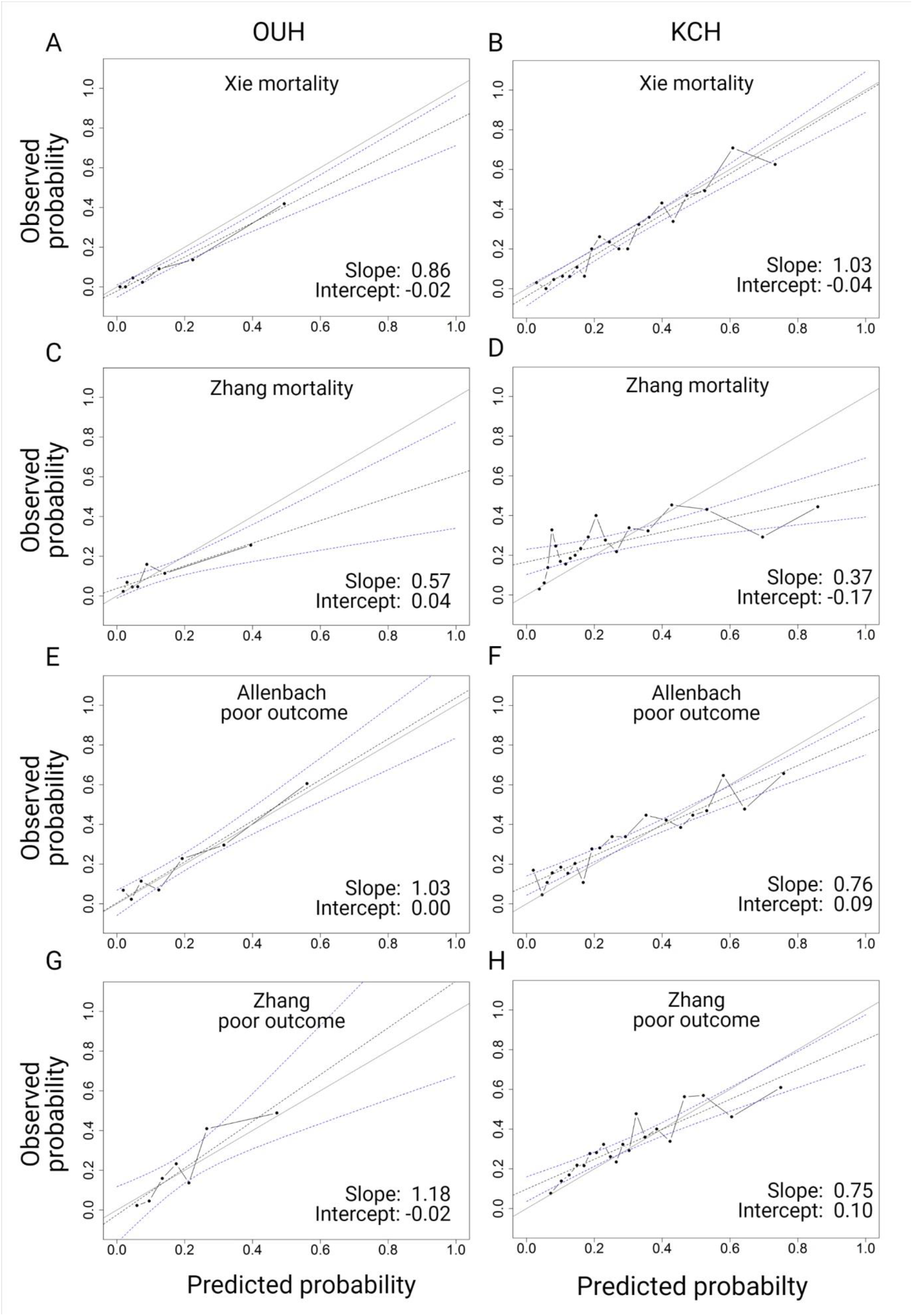
Calibration plots for OUH and KCH after recalibration.

For ‘severe disease’, discrimination was highest in the Allenbach model with AUROCs 0.72 (95 % CI 0.68-0.75) for KCH and 0.81 (95 % CI 0.74-0.88) for OUH. For the Zhang2 model, the AUROC was 0.67 (95 % CI 0.64-0.70) for KCH and 0.77 (95 % CI 0.70-0.84) for OUH. For the Xie and Allenbach models, discrimination at OUH was similar to the development cohorts (Figure 2). And, although the difference was not statistically significant at the 0.05 confidence level, we found better discrimination for both of these models at OUH compared to KCH.

The calibration plots are shown in Figure 3 (after recalibration). Figure S3 in supplementary shows the calibration results before and after recalibration for the Xie and Allenbach models.

Recalibration will not render models with poor discrimination more useful. Thus, we focused on the recalibration of the Xie and Allenbach models as these had the best discrimination. Recalibration

improved the predictions for both the Xie and Allenbach models at OUH and the Xie model at KCH, and the slope and intercept were acceptable for both models at both hospitals after recalibration.

## Discussion

In this study, we validated four prediction models for prognosis in hospitalized COVID-19 patients from London, UK and Oslo, Norway. We found varying performance of the models in the two cohorts. The models performed better in the OUH cohort with similar discrimination to the original studies. The Xie and Allenbach models had the best performance for prediction of death and severe disease, respectively.

Initial calibration was poor for all models, but improved after recalibration of the intercept according to the frequency of the outcome in our cohorts. This improves the accuracy of the prediction for each patient without affecting the discrimination and is recommended in several publications [5, 11, 18]. Local or possibly regional/national recalibration is likely to be important for COVID-19 prediction models since there is a large variation in the frequency of severe disease and death in different studies.

In some cases, we found poorer discrimination in the validation cohorts compared to the development cohorts. This is consistent with past evidence showing discrimination in development cohorts to be better than at external validation due to overfitting and differences in characteristics of the cohorts [23, 24]. The cohorts in the original studies and at KCH and OUH had many differences such as mortality, age and frequencies of severe disease and comorbidities. UK and Norway differ in the structures of their healthcare systems, and the incidence of COVID-19 has been far higher in the UK. These factors may have affected the selection of patients for hospital and ICU admission, which might have resulted in a more homogenous patient population in regards to severity at KCH. It is to be expected that discrimination will be less good when the population is more homogenous.

The findings underline the importance of validation at several external sites. This is particularly true for a new disease like COVID-19, with rapidly developing treatment guidelines, and with an overwhelming effect on healthcare resources in some locations, but not at others.

The Xie model had the best results compared to the other models. The differences in the performance of the prediction models might have several reasons. Firstly, the predictors used in one model might have better predictive value than predictors used in others. SaO_2_, which is included in the Xie model, is a strong clinical indicator of the severity of disease, and often indicates a need for ICU transfer. Secondly, there might be weaknesses in the models, as bias is common in prediction models [12]. To date, only the Allenbach study is published in a peer-reviewed journal, while Xie and Zhang are preprints. Thirdly, criteria for ICU admittance might vary across sites. The fact that we and other studies generally find better discrimination for mortality than for severe disease (often defined by ICU admittance) supports this hypothesis. For instance, patients with short life expectancy will often not be admitted to the ICU, but given oxygen therapy in a hospital ward and transferred to nursing homes for palliative care. These patients, not fulfilling the criteria for severe disease, often have predictors that indicate severe disease at admission.

Many prediction models have been published, but few have been systematically validated [24]. To our knowledge, only one study to date has validated COVID-19 prediction models; Gupta et. al recently validated 22 prognostic models [6], including the Xie and Xhang models. For the OUH cohort, we found substantially better discrimination for the Xie and Allenbach models for the prediction of mortality and severe disease, respectively. The performance of the models at KCH was more similar to the results in the Gupta study, also performed at a London hospital. The rate of severe disease, mortality and the characteristics of the London cohorts are quite similar which might explain the similar performance at these two sites.

Several other prediction models have been recently published, such as models based on NEWS2 or the ISARIC model [14, 25]. The AUROCs of the models are in the range of 0.75 to 0.80, which is not a substantial improvement over single univariate predictors of severity. Thus, the finding that the Xie and Allenbach models perform well at both the original study site and at our validation cohort at OUH might indicate that it is possible to achieve higher AUROCs with relatively simple prediction models.

Our study has several strengths. Validation was performed at two sites in different countries with consistent inclusion and exclusion criteria. We included all eligible patients admitted to the hospital during the study period therefore the cohorts should be representative of the study sites. Moreover, the study was conducted and reported according to the TRIPOD guidelines. However, there are also some weaknesses. Firstly, the OUH cohort is not very large with relatively few patients meeting the outcomes. Some publications recommend including at least 100 patients with the relevant outcome [10]. However, studies with lower numbers are frequently published, and may still contain useful information. Furthermore, the KCH cohort is probably one of the largest cohorts analyzed in prediction models for severe COVID-19. Secondly, Gupta et. al. included 22 models in their validation study, while we ended our inclusion of models in May, and included only four models in this study. Whereas it could be interesting to include more models we think that the results for the Xie and Allenbach models at OUH indicate that further studies of these models could be interesting. Thirdly, there was a relatively high number of missing values for LDH and SpO_2_ at KCH. It is uncertain how much this affected the results. Both are included in the Xie model and SpO_2_ is a strong predictor for mortality, while LDH is probably a weaker predictor (6). The number of missing values at OUH was low and probably did not affect the validation.

In conclusion, following the TRIPOD guidelines, our study validated developed models for prediction of prognosis in COVID-19, and showed that these models have a variable performance in different cohorts. The Xie model and Allenbach model clearly had the best performance, and we suggest that these models should be included in future studies of COVID-19 prediction models.

However, the performance of these models at our two validation sites was not similar, which underlines the importance of external validation of prediction models at several study sites before their implementation in the clinical practice.

## Supporting information

TRIPOD checklist

## Data Availability

Code and pre-trained models are available at https://github.com/ocbe.uio/covid-19 model validation and openly shared for testing in other COVID-19 datasets.
KCH: Source text from patient records used at all sites in the study will not be
available due to inability to safely fully anonymise up to the Information
Commissioner Office (ICO) standards and would be likely to contain strong
identifiers (e.g. names, postcodes) and highly sensitive data (e.g. diagnoses).
A subset of the KCH dataset limited to anonymisable information (e.g. only
SNOMED codes and aggregated demographics) is available on request to
researchers with suitable training in information governance and human
confidentiality protocols subject to approval by the Kings College Hospital
Information Governance committee; applications for research access should
be sent to. This dataset cannot be released
publicly due to the risk of re-identification of such granular individual-level
data, as determined by the Kings College Hospital Caldicott Guardian.
OUH: The OUH dataset cannot be released publicly due to the risk of reidentification of such granular individual-level data. Researchers can contact Erik Koldberg Amundsen for any enquiries

## Supporting information

**S1 File. Supplementary tables**

**S2 File. Supplementary figures**

**S2 File TRIPOD checklist**

## Acknowledgments

We would like to thank Prof. Anne Ma Dyrhol Riise and Dr. Ane M. Andersson at the Department of Infectious Diseases, OUH, for their support with the quality registry “COVID19 OUS”.

## Author contributions

### Conceptualization

Kristin Wickstrøm, Erik K. Amundsen, Kristian Tonby, Aleksander R. Holten, Valeria Vitelli, Alvaro Köhn-Luque

### Data Curation

Kristin Wickstrøm, Erik K. Amundsen, Kristian Tonby, Valeria Vitelli, Alvaro Köhn-Luque, Andrew H. Reiner, Ewan Carr, Rebecca Bendayan, Daniel Bean, Anthony Shek, Zeljko Kraljevic

### Formal Analysis

Valeria Vitelli, Alvaro Köhn-Luque, Andrew H. Reiner, Ewan Carr

### Methodology

Kristin Wickstrøm, Erik K. Amundsen, Kristian Tonby, Valeria Vitelli, Alvaro Köhn-Luque,

### Project Administration

Erik K. Amundsen, Kristian Tonby

### Resources

Erik K. Amundsen, James Teo, Richard Dobson

### Software

Valeria Vitelli, Alvaro Köhn-Luque, Andrew H. Reiner, Ewan Carr, Daniel Bean, Anthony Shek, Zeljko Kraljevic

### Supervision

Erik K. Amundsen, James Teo, Richard Dobson

### Validation

Kristin Wickstrøm, Valeria Vitelli, Alvaro Köhn-Luque, Andrew H. Reiner, Ewan Carr

### Visualization

Kristin Wickstrøm, Valeria Vitelli, Alvaro Köhn-Luque, Andrew H. Reiner, Erik K. Amundsen

### Writing (original draft)

Kristin Wickstrøm, Erik K. Amundsen

### Writing (Review)

Kristian Tonby, Aleksander R. Holten, Valeria Vitelli, Alvaro Köhn-Luque, Ewan Carr, Rebecca Bendayan, Daniel Bean, Anthony Shek, Zeljko Kraljevic, Tom Searle, James Teo, Richard Dobson

