## Supplemental tables for "Regional performance variation in external validation of four prediction models for severity of COVID-19 at hospital admission: An observational multi-centre cohort study"

### Supplementary tables

#### **Table S1:** Patient characteristics of the development/validation cohorts

| | Development cohorts | | | | | | Validation cohorts | | | |
|---|---|---|---|---|---|---|---|---|---|---|
| | Xie model (n=299) | | Allenbach model (n=152) | | Zhang model (n=775) | | KCH (n=1295) | | Oslo (n=307) | |
| **Outcome** | N avail | N (%) | N avail. | N (%) | N avail. | N (%) | N avail. | N (%) | N avail. | N (%) |
| Inhospital mortality | -- | 155 (51.8%) | -- | -- | 773 | 33 (4.3%) | 1244 | 333 (26.8%) | 307 | 32 (10.4%) |
| Mortality by day 14 | -- | -- | 146 | 32 (21.9%) | -- | -- | | | 307 | 21 (6.8%) |
| ICU/death (no time frame) | -- | -- | -- | -- | 773 | 75 (9.7%) | 1244 | 419 (33.7%) | 307 | 66 (21.5%) |
| ICU/death by 14 days | -- | -- | -- | -- | -- | -- | 1248 | 389 (31.2%) | 307 | 62 (20.2%) |
| ICU by day 14 | -- | -- | 146 | 17 (11.6%) | -- | -- | | | 307 | 49 (15.9%) |
| **Demographics** | N avail | N (%) | N avail. | N (%) | N avail. | N (%) | N avail. | N (%) | N avail. | N (%) |
| Age (median [IQR]) | -- | 65.0 [54.0-73.0] | -- | 77 [60-83] | 775 | 61 (50-68) | 1295 | 69.0 [56.0-81.9] | 307 | 60.0 [48.0-73.0] |
| Sex (male) | -- | 144 (48.2%) | 152 | 91 (59.9%) | 775 | 379 (48.9%) | 1295 | 767 (59%) | 307 | 175 (57.0%) |
| Non-White ethnicity | -- | -- | 140 | 50 (35.7%) | -- | -- | 961 | 418 (43%) | -- | -- |
| **Comorbidities** | N avail | N (%) | N avail. | N (%) | N avail. | N (%) | N avail. | N (%) | N avail. | N (%) |
| Hypertension | -- | 127 (42.6 %) | 152 | 82 (53.9%) | 771 | 239 (30.8%) | 1295 | 700 (54%) | 307 | 104 (34%) |
| Diabetes mellitus | -- | 55 (18.5%) | 152 | 37 (24.3%) | 771 | 106 (13.7%) | 1295 | 455 (35%) | 307 | 64 (21%) |
| Heart Failure | -- | 13 (4.4%) | -- | -- | -- | -- | 1295 | 96 (7%) | | |
| Heart disease | -- | -- | -- | -- | 771 | 85 (11.0%) | -- | -- | | -- |
| Ischaemic Heart Diseases | -- | -- | 152 | 35 (23%) | -- | -- | 1295 | 177 (14%) | 307 | 33 (11%) |
| Chronic lung disease | -- | -- | -- | -- | 771 | 48 (6.2%) | -- | -- | 307 | 83 (27%) |
| COPD | -- | 15 (5.0%) | 151 | 12 (7.9%) | -- | -- | 1295 | 137 (11%) | -- | -- |
| Chronic Kidney Disease | -- | -- | -- | -- | 771 | 25 (3.2%) | 1295 | 215 (17%) | 307 | 20 (7%) |
| ACE/A2 inhibitor | -- | -- | 152 | 28 (18.4%) | -- | -- | -- | -- | 307 | 62 (20%) |

| | | | | | | | | | | |
|---|---|---|---|---|---|---|---|---|---|---|
| Smoking | -- | 10 (3.4%) | 152 | 10 (6.6%) | -- | -- | -- | -- | 307 | 14 (5%) |
| **Blood biomarker** | N avail | Median (IQR) | N avail. | Median [IQR] | N avail. | Median [IQR] | N avail. | N (%) | N avail. | N (%) |
| Albumin (g/L) | -- | 33.4 [29.9-36.2] | -- | -- | -- | -- | -- | -- | 263 | 39.0 [37.0-42.0] |
| C-reactive protein (CRP, mg/L) | -- | 65.3 [27.4-113.9] | 152 | 74.5 [30.9-135.1] | 684 | 2.7 [0.9-14.1] | 1203 | 87.4 [41.0-154.3] | 307 | 44.0 [14.0-97.0] |
| Creatinine, U/L | -- | 74.0 [58.0-95.0] | -- | -- | 728 | 63.9 [54.1-76.4] | 1200 | 93.0 [71.8-133.2] | 307 | 80.0 [64.0-99.0] |
| Lymphocyte count (x10$^9$/L) | -- | 0.75 [0.5-1.11] | -- | * | 743 | 1.4 [1.1-1.9] | 1187 | 1.0 [0.7-1.3] | 295 | 1.1 [0.7-1.5] |
| Neutrophil count (x10$^9$/L) | -- | -- | 152 | 4.4 (3.0-7.0) | 739 | 3.5 (2.7-4.9) | 1186 | 5.6 [3.9-7.8] | 295 | 4.2 [3.1-6.3] |
| Platelet count (x10$^9$/L) | -- | 179.5 [137.3-247.8] | -- | -- | 732 | 224 [180-274] | 1188 | 212.0 [163.0-272.0] | 307 | 210.0 [165.0-269.0] |
| LDH (U/L) | -- | 384.0 [264.0-541.0] | -- | 364 [284-444] | -- | -- | 157 | 405.0 [307.0-553.0] | 269 | 255.0 [191.5-328.5] |
| **Physiological parameters** | N avail | Median (%) | N avail. | Median (%) | N avail. | Median (%) | N avail. | N (%) | N avail. | N (%) |
| Oxygen saturation | -- | 95.0 [90.0-98.0] | 152 | 93 (90-96)[1] | -- | -- | 858 | 96.0 [95.0-98.0] | 307 | 95.0 [92.0-97.0] |
| Oxygen flow rate (L/min) | -- | -- | 152 | 2 (2-4) | -- | -- | 880 | 1.0 [0.0-4.0] | 220 | 0.0 [0.0-2.0] |
| Systolic blood pressure | -- | 132.5 [118.0-145.0] | -- | -- | -- | -- | 862 | 124.0 [112.0-139.0] | 306 | 127 [116.0-141.0] |
| **Symptoms** | N avail | N (%) | N avail | N (%) | N avail | N (%) | N avail. | N (%) | N avail. | N (%) |
| Fever | -- | -- | -- | -- | 773 | 532 (68.6%) | -- | -- | 307 | 194 (63.2%) |
| Cough | -- | -- | -- | * | 773 | 528 (68.1%) | -- | -- | 307 | 218 (71.0%) |
| Fatigue | -- | -- | 150 | 70 (46.7%) | 773 | 421 (54.3%) | -- | -- | 307 | 109 (35.5%) |
| Dyspnoe | -- | -- | 150 | 102 (67.5%) | 773 | 355 (45.8%) | -- | -- | 307 | 202 (65.8%) |
| | | | | | | | -- | -- | | |

[1] *Measured without oxygen*

* *other definition of this variable*

Table S2: OUH cohort AUROC results show similarities with different imputation methods.
KNN; K-nearest neighbor, RF; Random forrest, GP; Gaussian Process Method, BR; Bayesian Rigde.

| | Xie model | Allenbach | Zhang1 | Zhang2 |
|---|---|---|---|---|
| Single imputation; KNN | 0.87 [0.79-0.95] | 0.81 [0.74-0.88] | 0.72 [0.62-0.82] | 0.77 [0.70-0.84] |
| Single imputation; RF | 0.86 [0.78-0.94] | 0.81 [0.74-0.88] | 0.72 [0.62-0.82] | 0.77 [0.70-0.84] |
| Multiple imputation; BR | 0.86 [0.78-0.95] | 0.81 [0.74-0.88] | 0.72 [0.62-0.82] | 0.77 [0.70-0.84] |
| Multiple imputation; GP | 0.85 [0.76-0.95] | 0.80 [0.73-0.87] | 0.72 [0.62-0.82] | 0.77 [0.70-0.84] |
