## Supplemental figures for "Regional performance variation in external validation of four prediction models for severity of COVID-19 at hospital admission: An observational multi-centre cohort study"

### Supplementary Figures

**Figure S1: Flowchart of the included patients at OUH for validation of the four prediction models**

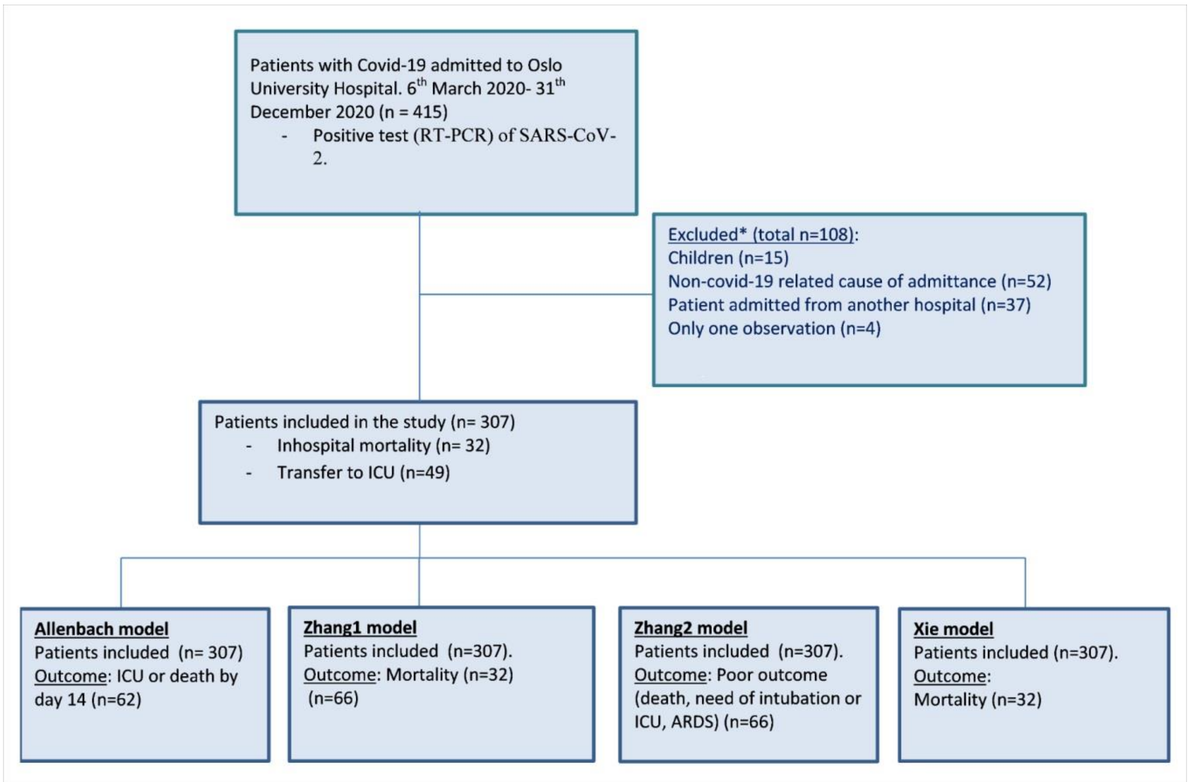

**Figure S2: Flowchart of included patientes in the KCH cohort for validation of the four predcition models**

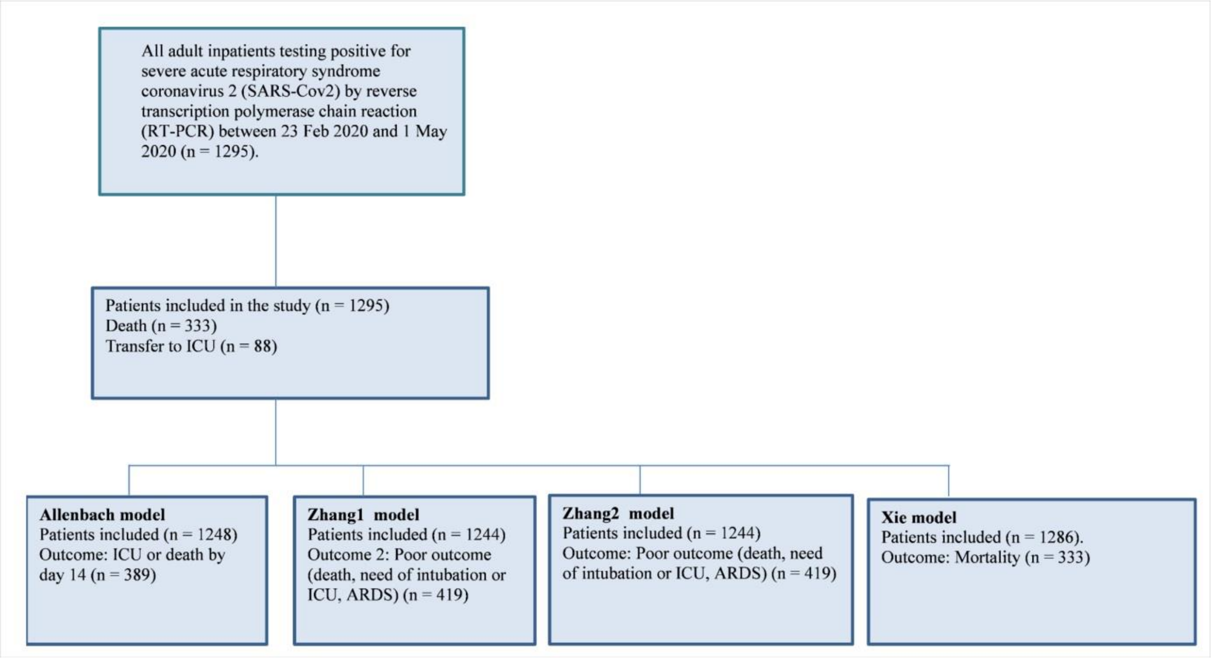

**Figure S3: Results for calibration of the Xie and Allenbach models at KCH and OUH before and after recalibration**

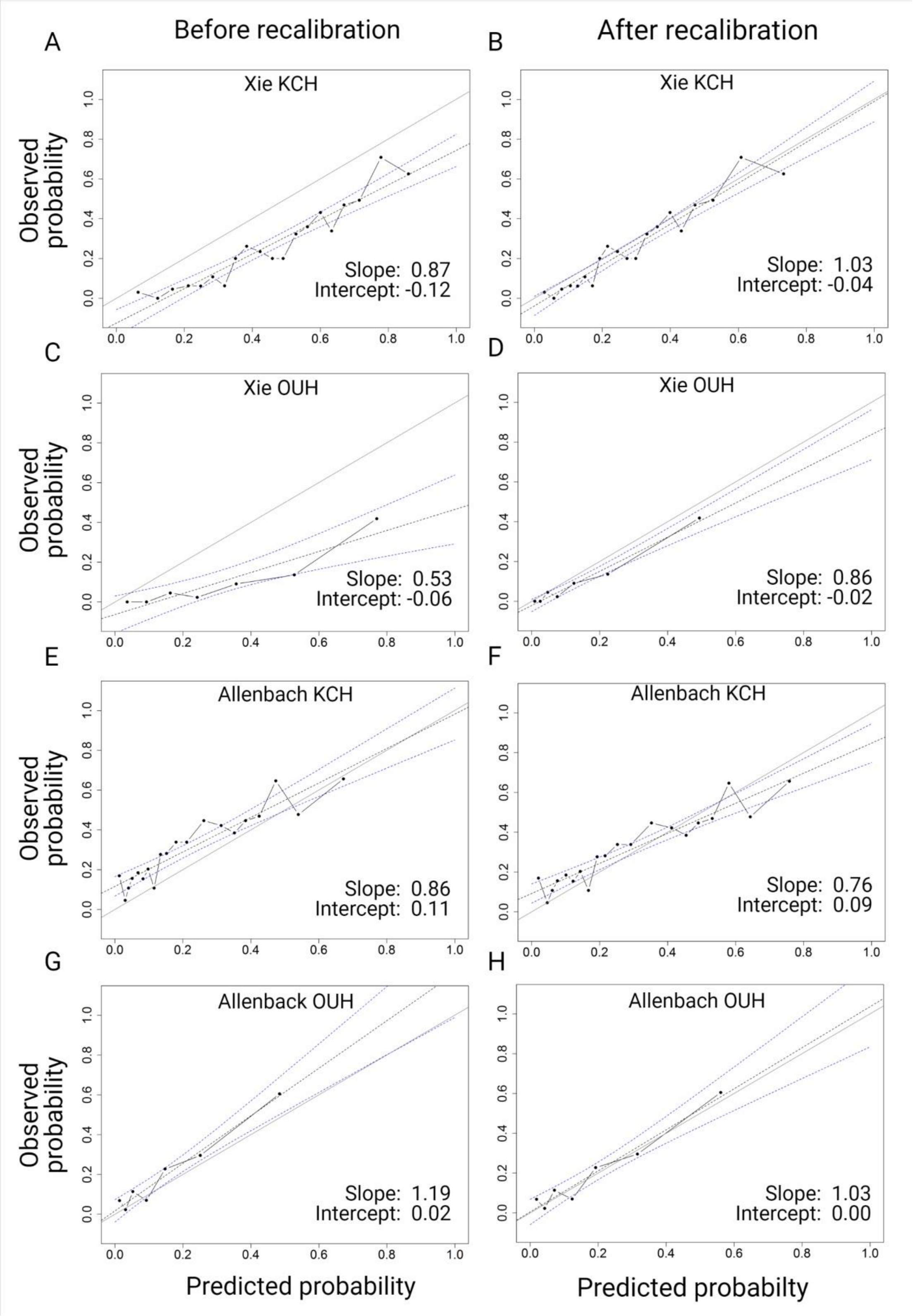
